# Cost-effectiveness of Intraoperative Fluorescence Angiography to Prevent Anastomotic Leak in Rectal Cancer Surgery: Economic Evaluation alongside the IntAct Randomised Controlled Trial

**DOI:** 10.1101/2025.10.03.25337241

**Authors:** David Meads, Armando Vargas-Palacios, Neil Corrigan, Gemma Ainsworth, Julie Croft, Ronan Cahill, Kathryn Gordon, Andrew Kirby, Albert Wolthuis, Roel Hompes, Deborah D Stocken, David Jayne

## Abstract

**Background:** Anastomotic leakage (AL) following resection for rectal cancer has a significant negative impact on patients and represents a substantial economic burden. Intraoperative fluorescence angiography using indocyanine green (ICG+) is a potential strategy to reduce ALs. Findings from recent randomized controlled trials were positive; however, no full economic evaluations of ICG+ have been conducted to date. We conducted a cost-utility analysis of the ICG+ vs standard surgeon assessment (ICG-) using the IntAct trial data.

**Methods:** We took the perspective of the UK NHS over a 90-day post procedure horizon. EQ-5D-5L and resource use data were collected at baseline, 30 and 90 days in the UK trial sub-sample to enable estimation of quality-adjusted life years (QALYs) and costs. Incremental cost-effectiveness ratios (ICERs) were estimated using generalised linear regression models.

**Results:** We analysed data from n=345 UK patients finding negligible adjusted differences in QALYs (−0.001 in the primary multiply imputed analysis) at 90 days but modest cost savings (£73 [95% CI −£78, −£67] for the primary and £140 [95% CI −£150, −£129] for the secondary complete case analyses) per patient for ICG+. The ICER primary analysis indicated ICG+ was cost effective. Simulations indicated ICG+ had a 60% chance of being the optimal strategy in the primary analysis.

**Conclusions:** This research represents a novel contribution on the value of ICG+ for preventing ALs. Results indicate that ICG+ leads to modest cost savings. Together with the clinical effectiveness results, and the potential positive budget impact, the current results indicate ICG+ is likely to provide net health benefit.

## Introduction

Anastomotic leakages (ALs) are a common complication of rectal cancer surgery, occurring on average in 9% [1] of procedures but often at much higher rates. ALs are associated with increased morbidity and mortality, poorer oncological outcomes and a reduction of patients’ quality of life. The economic burden of ALs is substantial; according to one review, each AL leads to between €2,250-€83,633 of additional healthcare costs depending on the study country and context.[2] The additional costs associated with ALs are typically related to increased hospital length of stay, intensive care, readmissions and re-interventions.

There are many patient and clinical factors that are predictive of AL. A critical and modifiable factor is blood supply to the anastomosis. Historically, perfusion has been assessed using standard assessment by the surgeon (using white light imaging). However, intra-operative fluorescence angiography (IFA) is now seen as an adjunct to standard of care. The IFA approach uses intravenous administration of indocyanine green (ICG) and near-infrared light to provide an augmented assessment of tissue perfusion and to enable better informed clinical decision-making during surgery. While many studies have shown the potential benefit of ICG+, none have included formal cost-effectiveness analysis.

The IntAct trial[3][4] was a European, multi-centre, unblinded, parallel group, randomized controlled trial comparing surgery with ICG (ICG+) against the standard care of white light laparoscopy (ICG-). Patients (n=766) undergoing elective anterior resection for rectal cancer were recruited from 28 European sites across 8 countries. The primary outcome measure was rate of clinical (International Study Group of Rectal Cancer[5], grades B/C) AL within 90 days of surgery. Secondary outcomes included all AL (clinical and radiological; grades A-C), complications and re-interventions, stoma use and quality of life (QoL). Given the economic burden of AL and the growing pressures on health care services, cost effectiveness was also a key secondary endpoint in IntAct.

We conducted an economic evaluation of ICG+ vs ICG-from a UK health and personal social services perspective. The analysis was alongside the clinical trial and hence the time horizon was 90 days post-procedure.

## Methods

We adopted the evaluation reference case outlined by the National Institute for Health and Care Excellence (NICE)[6]. As such, we used a cost-utility framework to answer the research question, presenting cost-per quality-adjusted life year (QALY) gained. We also present a cost-effectiveness analysis based on cost per AL avoided.

As described above, the IntAct study included UK and non-UK sites. Patients in the UK completed both health-related quality-of-life (HRQoL) assessments and health resource use questionnaires, while non-UK patients only completed the HRQoL questionnaires. As we could not directly estimate full costs (including primary and social care and medications) for the non-UK sites, the economic evaluation only considered UK patients (348/766 patients).

### Resource use

The following items of health care resource use were captured during the trial for the purposes of estimating costs:

- Health and social care resources used after the operation or a complication from surgery or the use of ICG+
  - Inpatient stay (Length of stay)
  - Outpatient care (e.g. clinic visits, routine follow-up)
  - Interventions required to treat an AL: e.g. surgical, pharmacological, radiological, endoscopic
  - Personal social services support (e.g. prepared meals, laundry services, care workers, social workers)
  - Community and residential based health and social services (e.g. GP visits and contacts, district nurses, physiotherapists, nursing home stays)
  - Medications (free text question)

This information was captured using a combination of trial case report forms (CRFs) and patient-reported resource use forms. The latter were collected at baseline, 30 and 90 days post-operative procedure. The primary admission cost includes the costs of initial surgery and any complication (e.g. leak, stoma), conversion costs and all associated length of stay. They also include the ICG+ cost (£104 for the drug and £44 per use of the imaging platform – see supplementary material for details).

All resource use was valued in monetary terms using appropriate UK unit costs or participant valuations estimated at the time of analysis (2023/24). Adjustments were made for inflation using the Office for National Statistics (ONS) Gross Domestic Product (GDP) health index [7] accordingly. NHS reference costs[8] and Personal Social Services Research Unit (PSSRU) costs of health and social care[9] were used to value resource use such as GP, Nurse, Physiotherapist visits, inpatient and outpatient attendances. Medication costs were taken from the British National Formulary (BNF[10]) and the Prescription Cost Analysis (PCA[11]) for England. Other Unit costs were identified online and in published literature. The costing strategy is outlined in supplementary material and uses NHS Health Resource Group (HRG) cost tariffs and length of stay trim points. NHS HRG costs are clinically meaningful groups of hospital activity based on patient records that include an average length of stay (trim point). Hospital stays beyond this are costed as excess bed days. We conducted a sensitivity analysis around this where we disaggregate procedure and length of stay cost.

### Health-related quality of life (HRQoL)

The primary economic outcome measure was Quality-Adjusted Life Years (QALYs) derived from utility scores, combined with life years, at 90 days post-procedure. Utility was measured using the EQ-5D-5L [12] HRQoL instrument at baseline, 30 and 90 days post procedure. The EQ-5D-5L was also completed at 12 months and this data was used in supplementary analysis. We also used the same primary endpoint as the trial: clinical (grades B/C) AL within 90 days of surgery for a separate cost-effectiveness analysis.

UK utility values were derived using the approach recommended by NICE, which is currently based on the validated mapping function to the existing EQ-5D-3L[13]. These were used to generate QALYs over the 90-day period taking an area under the curve approach, adjusting for any imbalances in baseline EQ-5D-5L scores. We also evaluated QALYs at 12 months post-operative procedure for those patients who completed this questionnaire.

Additionally, in a sensitivity analysis, we used the EORTC QLQ-C30 responses to derive the QLU-C10D[14] cancer-specific utility index to estimate QALYs.

### Missing data

Our primary approach for handling missing data was multiple imputation. We used Multiple Imputation by Chained Equations (MICE) to replace missing cost and utility scores. Predictors in the MICE model were pre-specified trial stratification factors and baseline cost and HRQoL values. Multiple estimates (n=30) of utility and costs were combined according to Rubin’s rules[15].

### Analysis

The intention to treat (ITT) population of eligible and randomised patients were the primary sample in the analysis (n= 345). A per protocol (PP) analysis included all participants in the full analysis set who were deemed to have no major protocol violations (e.g. not undergoing surgery or treatment strategy as per randomisation, or breaches of eligibility; n=34).

We estimated costs and benefits in separate models after controlling for covariates; these values entered the cost-effectiveness calculation. The models account for skewness in the data (i.e. generalised linear model for costs, Family: Gamma; Link: Log) and we use mixed effects models (on surgeon) to account for the multi-level nature of the data.

The covariates used in the model reflect any imbalances between groups at baseline, including (baseline) costs or utility scores, as well as those covariates (stratification factors) used in the main statistical efficacy analysis:

- Participant gender (male/female);
- ASA grade (I, II or III);
- Radiological T-stage of the rectal cancer (T1, T2, T3, T4);
- Neo-adjuvant therapy received (none, short course with no delay, short course with delay or long course);
- Position of the rectal cancer tumour (above, at or below the peritoneal reflection);
- Intended operating surgeon (fitted as a random effect).

We estimated the incremental cost-effectiveness ratio (ICER) which is the difference in costs, divided by the difference in effects between the two trial arms. Should the ICER be <£20,000 per QALY gained, ICG+ can be considered a cost-effective use of NHS resources and is suitable for routine commissioning. The primary analysis was the ITT, adjusted, multiply imputed analysis. A secondary, complete case analysis considered data only from patients who had completed EQ-5D-5L and resource use assessments at all time-points. We permitted patients to miss one EQ-5D-5L completion in the QALY estimate. Additional sensitivity analyses are reported. We also converted the cost and QALY information into the Health Benefit metric. Net Health Benefit (NHB) is calculated as follows:

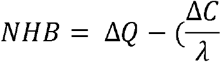

Where ΔQ is incremental QALYs, ΔC is incremental costs and ⍰ is the willingness to pay threshold (£20,000). Health benefit at 90 days is estimated at an individual basis and used as the dependent variable in multilevel mixed effects generalised linear regression models using the stated covariates and accounting for clustering at surgeon level. Should the dependent variable coefficient denoting randomisation be positive, then cost-effectiveness is indicated with uncertainty subsequently characterised. NHB regression was used to facilitate sub-group analysis through the use of interactions with randomisation.

We used random draws from the variance-covariance matrices to determine the level of sampling uncertainty surrounding the ICER calculated in the primary analysis. We did this to generate 10,000 estimates of incremental costs and benefits which were plotted on the cost effectiveness plane. Results were also presented as cost-effectiveness acceptability curves (CEACs). The base case ICER used the mean costs and effects from the estimates.

We also explored the impact of ALs on surgical resource use (time in theatre, initial length of stay), costs (using a GLM model), HRQoL, QALYs and NHB in supplementary linear (except where stated) regression analyses using data pooled across trial arms.

Costs and outcomes were not discounted as the time horizon for the analysis was less than 12 months. All analyses were conducted using Stata version 18.

## Results

Of the 348 UK patients, 345 were included in the primary final analysis following multiple imputation, 93 patients constituted the secondary complete case analysis. The final sample, recruited across 18 UK sites were 31% female and had a mean age of 64 (range =30-89). In total in the evaluable UK sample (n=307), 10% and 19% of patients experienced a clinical or any grade AL, respectively. The equivalent adjusted odds ratios comparing ICG+ to ICG-were 1.13 (95%: 0.54-2.8) and 0.75 (95%: 0.42-1.34), respectively. Table 1 includes the mean per patient costs, per trial arm over 90 days for the multiple imputation and complete case analyses. Table 2 includes the mean QALYs over 30 and 90 days for the same analyses.

**Table 1:** Total and category costs at 90 days.

| Costs* | Imputed Costs at day 90 |  | Complete case Costs at 90 days |  |
| --- | --- | --- | --- | --- |
|  | ICG-<br>(SE) N= 170 | ICG+<br>(SE) N = 175 | ICG-<br>(N, SE) | ICG+<br>(N, SE) |
| **Total costs** | £11,867<br>(377.75) | £11,527<br>(377.00) | £12,434<br>(141, 189.20) | £11,626<br>(144, 173.94) |
| **Primary admission including surgery\*\*** | £9,471<br>(261.36) | £8,950<br>(240.62) | £7,789<br>(171, 116.19) | £7,632<br>(172, 102.62) |
| **Readmission hospital costs\*\*\*** | £688<br>(129.08) | £729<br>(140.99) | £795<br>(81, 163.37) | £715<br>(71, 219.22) |
| **Outpatient** | £923<br>(96.25) | £911<br>(76.23) | £958<br>(83, 139.63) | £833<br>(81, 91.21) |
| **Primary care** | £141<br>(14.28) | £127<br>(15.74) | £121<br>(82, 14.73) | £132<br>(81, 22.80) |
| **Treatment of an AL** | £397<br>(138.77) | £562<br>(160.03) | £397<br>(172, 138.38) | £562<br>(176, 160.03) |
| **Diagnosis of AL** | £118<br>(38.59) | £153<br>(60.32) | £118<br>(172, 38.59) | £153<br>(176, 60.32) |
| **Medication costs** | £103<br>(28.26) | £80<br>(11.29) | £115<br>(121, 35.52) | £90<br>(143, 12.73) |

\*Only total costs are adjusted
\*\*Includes primary admission stay costs, initial surgery costs, IFA costs, stoma maintenance, removal, and excess bed days due to complications after initial surgery or prolonged hospitalisation
\*\*\*Hospitalisation after first discharged for any reason including complications or re-intervention

**Table 2:**
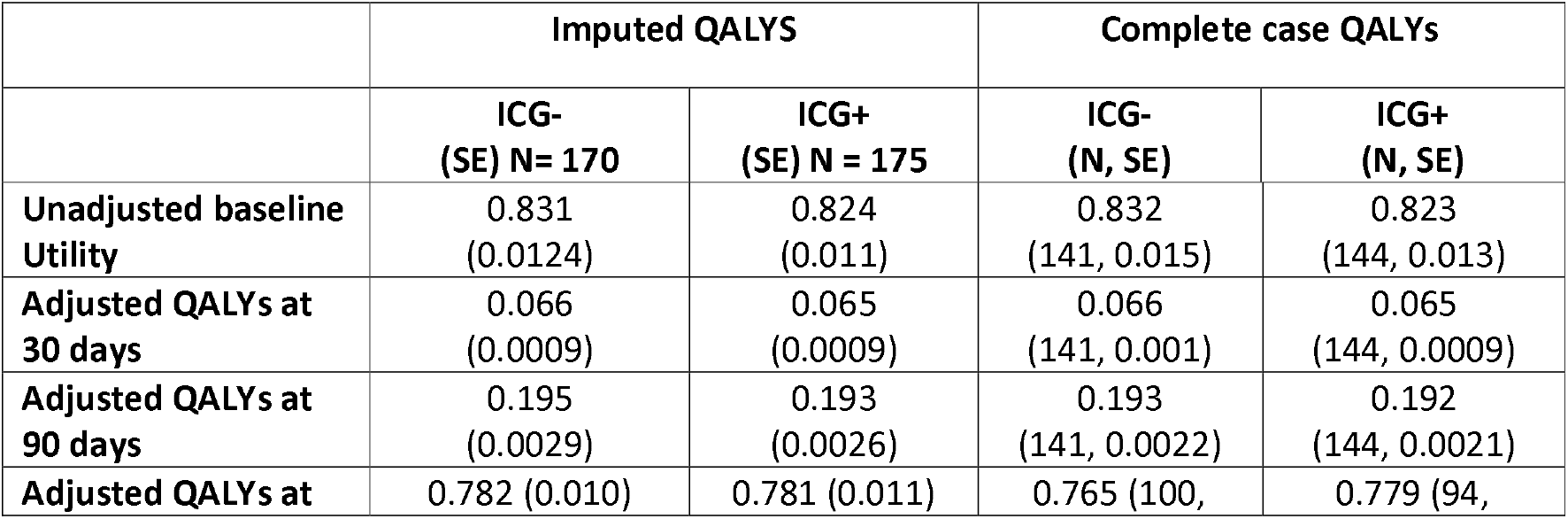
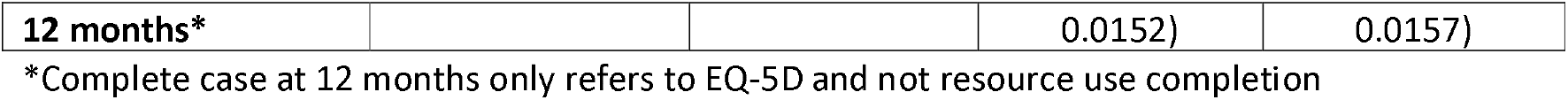
QALYs at 30 days,90 days and 12 months.

There were relatively small cost differences across arms (Table 1). However, ICG+ incurred noticeably lower primary admission and surgery costs and this drove the modest cost savings for ICG+ overall. There was a very small QALY difference (around 10 hours of full health) in favour of ICG-(Table 2). The base case cost-utility results are presented in Table 3. Given that ICG+ generates cost-savings for a very small QALY loss, the ICER represents costs saved per QALY lost. Hence, in this analysis (and not accounting for the variance in outcomes), the NHS would in theory save over £77,000 per QALY lost in the base case. The decision rule in such a scenario is that ICERs above £20,000 indicate cost-effectiveness. The incremental NHB is a little easier to interpret and, as it is positive, indicates ICG+ is cost-effective since the cost savings outweigh the small QALY loss in terms of value. Based on the 10,000 simulations to assess uncertainty, ICG+ has a 60% chance of being cost-effective.

**Table 3:** Base case and sensitivity analysis cost-utility results.

| Analysis | N | Incremental costs for ICG+ (95% CI) | Incremental QALYs for ICG+ (95% CI) | ICER | Incremental NHB | Prob. cost-effective |
| --- | --- | --- | --- | --- | --- | --- |
| **Base case** |  |  |  |  |  |  |
| Multiple imputation | 345 | -£73<br>(-£78, -£67) | -0.001<br>(-0.001, -0.001) | £77,383^ | 0.003 | 60% |
| **Sensitivity and supplementary analyses** |  |  |  |  |  |  |
| Complete case | 93 | -£140<br>(-£150, -£129) | 0.001<br>(0.006 - 0.008) | ICG+ dominates | 0.008 | 64% |
| Complete case, mean imputed missing 30 day costs | 178 | -£381<br>(-£391, -£371) | 0.001<br>(0.001,0.001) | ICG+ dominates | 0.020 | 83% |
| Per protocol analysis with multiple imputation | 311 | -75<br>(-£81, -£70) | -0.001<br>(-0.001, -0.001) | £59,817^ | 0.003 | 60% |
| Multiple imputation after mean imputation of missing 30 day costs | 345 | -£80<br>(-£85, -£74) | -0.001<br>(-0.001, -0.001) | £126,222^ | 0.003 | 61% |
| Complete case, using disaggregated procedure and LoS costs. | 94 | -£239<br>(-£252, -£227) | 0.001<br>(0.001,0.001) | ICG+ dominates | 0.013 | 65% |
| Multiple imputation, QLU-C10D QALYs | 345 | -£28<br>(-£35, -£21) | -0.007<br>(-0.007, -0.006) | £4,212^ | -0.005 | 35% |
| Separate estimates of costs and QALYs – multiple imputation* | 345 | -£156<br>(-£1063, £751) | -0.001<br>(-0.007, 0.005) | £1,735,889^ | 0.008 | N/A |
| Separate estimates of costs and QALYs – complete case* | 93 | -£364<br>(-£2425, £1697) | 0.003<br>(-0.006, 0.012) | ICG+ dominates | 0.021 | N/A |

\*All analyses except these estimate ICERs based on mean of 10,000 simulations; ^ICER indicates cost savings per QALY lost

Figure 1 shows the cost-effectiveness plane which is the scatterplot of 10,000 incremental costs and QALY pairs simulated using the variance around the mean incremental cost and effects and which characterises the uncertainty. Figure S1 shows the cost-effectiveness acceptability curve (CEAC) based on these simulations. The declining curve indicates that at higher threshold values, the probability that ICG+ is cost-effective decreases.

**Figure 1:**
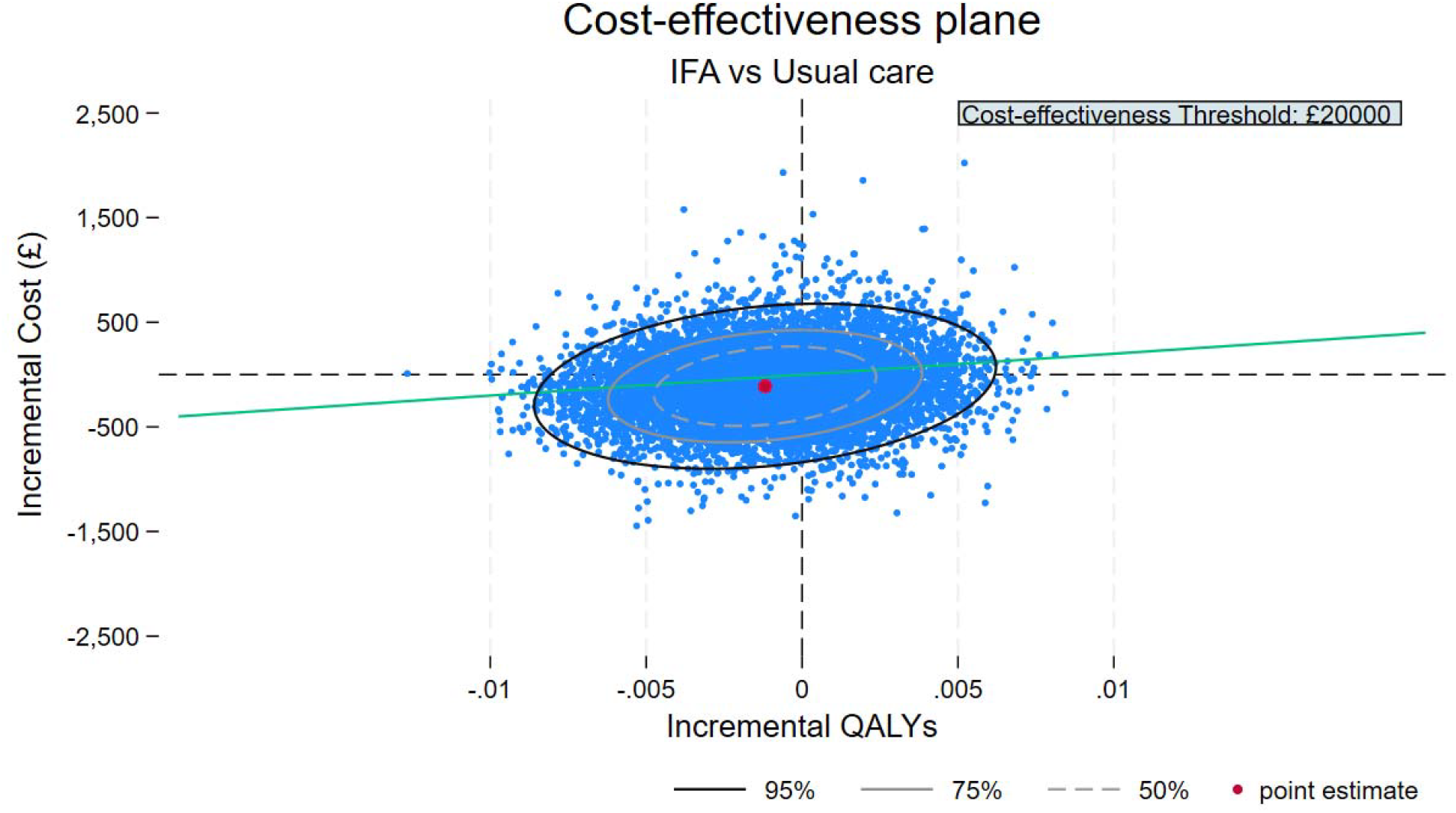
cost-effectiveness plane.

### Sensitivity and supplementary analysis

The complete case analysis (Table 3) yielded similar results to the primary analysis. However, the cost savings associated with ICG+ were slightly greater and ICG+ provided a QALY gain over ICG-. Hence, ICG+ is dominant in the complete case analysis. While a QALY gain is indicated, given the magnitude (approximately 6 hours of full health) and uncertainty surrounding it, the QALYs for ICG+ and ICG-should be considered to be practically the same at 90 days. Analysis of QALYs at 12 months (n=194) indicated an adjusted QALY gain of 0.015 (95% CI: −0.026, 0.056) for ICG+ in the complete case scenario. However, this difference does not extend to the multiple imputation analysis (see Table 2).

There were no statistically significant interactions (at p<0.01) between randomisation and stratification factors in predicting NHB suggesting an absence of sub-group effects. A cost-effectiveness analysis estimating cost per leak avoided was not possible as, in both primary and complete case analyses, ICG+ led to a cost savings and estimation of a meaningful cost per leak avoided requires a positive incremental cost.

### Impact of ALs on costs and HRQoL

Pooling the trial data across arms enabled the exploration of the impact of ALs (any grade n=39) on HRQoL and costs after adjusting for stratification factors without the need for imputation. In general, there was a relatively consistent, statistically significant impact of ALs on costs, EQ-5D utility values and QALYs. Results indicated that on average, ALs added £4,330 (p=0.001) of costs and reduced QALYs by 0.013 (p=0.023; equivalent to 4.75 days of full health) at 90 days. Full results are shown in table S4.

## Discussion

This study reports on an economic evaluation of indocyanine green (ICG+) perfusion angiography versus standard white light (ICG-) during rectal cancer resection. We found that the use of ICG+ was associated with modest cost savings. Depending on the analysis conducted, ICG+ was associated with either a very small QALY loss or gain. The results are robust to sensitivity analyses.

To the authors’ knowledge, this is the first full economic evaluation of ICG+ in the rectal cancer population. One study taking a Canadian health system perspective, conducted a cost analysis of ICG+ for the prevention of ALs in colorectal surgery.[16] They used a decision tree modelling approach to determine the cost impact of a reduction in ALs from 8.6% (ICG-) to 4.8% (ICG+) in the short term and assuming a cost of leak of $9,934.50 (Can$ 2014). They estimated a cost saving of $192.22 per case which (inflated and converted to £141.28) is similar to the cost savings we observed in this evaluation. Other economic evaluations of ICG+ in breast cancer[17] and noncancerous gallbladder[18] disease indicate ICG+ is cost saving and relatively cheap to implement.

Our analysis of the trial UK data sub-sample which included longitudinal HRQoL data over 12 months, suggests that the use of ICG+ did not have a material impact on quality-adjusted life years (survival or HRQoL) at 90 days. This was in line with the main statistical paper which found, despite the reduction in ALs, that there was no associated improvement in quality of life. It is not clear quite why this would be the case; there are several possible explanations including the short follow-up period (90 days) and the ALs being well managed.

It is also worth noting the relatively low rates of clinical ALs in general in the IntAct trial (12.9%)[4]; it is possible any HRQoL impact may have been washed out in the overall sample averages. We observed a non-trivial QALY gain at 12 months for the ICG+ arm (mean = 0.015 (5.5 days of full health; 95% CI = −0.026, 0.056) which was not statistically significant and not maintained after imputation of missing HRQoL data.

A robust finding was that ICG+ on average led to modest cost savings per patient in the range of £73-£381. Although we found the cost savings to be relatively small, when scaled up to the eligible population for England[19] (18,747 in 2022), these cost savings could be £1.8M and £9M over 1 year and 5 years, respectively. Perhaps more important, given existing and increasing pressures on health care systems, is the reduction in resource use that is the source of these cost savings. Cost savings in the ICG+ arm may have been reduced by the tendency for more permanent stomas in that arm which may have been a precautionary strategy.

Cost savings were observed across the primary and sensitivity analyses, however, there was substantial uncertainty around the estimates. There were no differences in length of stay (LoS) observed between treatment strategies. However, the observed cost savings may be explained by small reductions in ALs, post-operative complications and re-interventions in the ICG+ arm. The uncertainty in the cost-effectiveness of ICG+ is best reflected in the CEAC and probability of cost-effectiveness which was 60% in the base case.

We conducted further analyses which re-emphasise the impact of ALs on costs and HRQoL. We found that AL was associated with an average increase in theatre time of >41 minutes (95% CI: 8.86-74.81) and 7.85 days (95% CI: 7.47-10.31) in length of hospital stay. The average incremental cost of any AL was £4,330 (95% CI: £1684-£6976), more than an 80% increase on the cost of an uncomplicated surgery. However, this is substantially lower than that reported by several other published studies[2] including a UK study[20] which estimated incremental costs per AL to be £21,592 (inflated from £17,220, 2012 prices). When we compared costs for no AL/grade A vs grade B/C AL’s, the cost difference was increased (to £6,179) but not to the extent observed elsewhere which may be explained by the resources used to treat ALs. There was a notable impact of ALs on HRQoL and QALYs which extends to 12 months and this impact is accentuated when we only consider AL grades B and C.

The analysis was hindered by missing data at the 90 days follow-up. However, our findings do not change between complete case and multiple imputation analyses suggesting missingness does not unduly affect the recommendations. Data to enable a full economic evaluation was only available in the UK since resource use was not captured in the non-UK sites. This meant that a smaller dataset was useable and somewhat limits generalisability of results compared to the full EU sample where there was also stronger evidence of leak prevention for ICG+. Resource use is also limited to 90 days which limits the time horizon of the analysis. Future analysis may seek to model costs and benefits over a longer time horizon, incorporate data from the non-UK sites and evidence from other trials or meta-analyses.

We did not have sufficient information to take a bottom-up costing approach and micro-cost surgical procedures in the trial and thus had to rely on national procedure tariffs and length of stay to estimate costs. In doing so there may have been some double counting or omission of costs and such a loss of granularity may have affected cost estimates. For example, time in theatre and staff mix/numbers may have been impacted by the use of ICG+ and this is not captured in the tariff costing approach. For the platform cost of ICG+, we had to rely on published estimates which introduces some uncertainty in the analysis. Future research should seek to obtain accurate costs from hospitals or prices from providing companies. New laparoscopic systems now incorporate ICG+ as standard thus the cost of ICG+ per case is relatively small; however, given the modest cost savings observed, outcomes will likely be sensitive to the cost of the technology.

## Conclusions

This research represents a novel contribution to the literature on the value of ICG+ for preventing ALs. The results were relatively consistent across analytical approaches in indicating that ICG+ led to a modest cost saving vs ICG-. Notwithstanding substantial uncertainty around the estimates, taken in tandem with the clinical effectiveness results and the potential positive budget impact, the current results suggest the use of ICG+ is likely to provide net health benefit.

## Data Availability

All data produced in the present study are available upon reasonable request to the authors

## Notes

**Declaration of interests:** RC receives funding support from Diagnostic Green, Stryker Corp, Johnson & Johnson, EU Horizon Europe, Arthrex, Astellas, and Medtronic. RH receives funding support from Stryker Corp..

**Data availability:** Individual participant data (with any relevant supporting material, e.g. data dictionary, protocol, analysis plan) for all trial participants (excluding any trial-specific participant optouts) will be made available upon reasonable request for secondary research purposes when all key analyses are complete & published.

### Competing Interest Statement

The authors have declared no competing interest.

### Clinical Trial

ISRCTN:13334746

### Funding Statement

This study was funded by the National Institute for Health and Care Research (NIHR) Efficacy and Mechanism Evaluation (EME) Programme (14/150/62).

### Author Declarations

Ethics committee of IRAS North West - Preston Research Ethics - gave ethical approval for this work. Ethics reference number: 17/NW/0193

