## Supplemental material for "Cost-effectiveness of Intraoperative Fluorescence Angiography to Prevent Anastomotic Leak in Rectal Cancer Surgery: Economic Evaluation alongside the IntAct Randomised Controlled Trial"

**Supplementary Material**

The approach to costing procedures and hospital length of stay (LoS) was as follows:

1. We used an NHS Cost Collection HRG average cost for elective, long-stay Malignant Gastrointestinal Tract Disorders with Single Intervention (with 0-2 complications and comorbidities[CC]) as the basic procedure cost for patients without any complications, conversions or stoma.

For patients experiencing a complication or under-going conversion, we used a higher cost category (3-5 CC).

For patients receiving a stoma, a higher cost HRG was used indicating multiple interventions (Malignant Gastrointestinal Tract Disorders with Multiple Interventions (0-2 CC). If the patient experienced complications or conversion then a higher sub-category was used (3-5 CC).

2. Each HRG cost code indicates an expected number of bed days (termed trim points) which generally increase with severity of condition and required intervention resources. We used NHS trim points for the each of the HRG codes (e.g. 10 trim-point days for the single procedure without any complications).
3. We estimated total length of stay (LoS) as the difference between admission and discharge.
4. If LoS was longer than the trim point indicated for their cost code, additional day costs were charged at the excess bed day cost (£331.99 per day).
5. Costs of ICG (indocyanine green [£104.00](1) and a proportion of the ICG platform costs [£44.80])(2) were applied to the ICG+ arm.
6. If the initial surgery used a robot, an additional cost multiplier was applied to the starting HRG cost (multiplier = 1.13 based on the ROLARR trial (3)).
7. Patients who had a stoma also incurred a monthly stoma management cost (£165.40). Stoma removal costs were applied where necessary.
8. If a patient experienced an AL or reintervention, we applied separate HRG costs to account for the treatment of these.
9. If the AL or reintervention occurred during the initial admission period, we reduced the excess bed days to account for multiple interventions (and therefore >1 trim-points).
10. We added to these surgery and intervention costs, any primary, community and secondary health care resource use and medication costs reported by the patient post-discharge.
11. If an AL or reintervention occurred after discharge, we accounted for this when costing the patient reported hospital admissions to avoid double counting.

This approach to costing surgery is illustrated in Table S1 which shows exemplar patient surgery and LoS costs. The full Unit costs (prices in £ sterling, 2024 prices) and assumptions are included in Table S2.

Table S1: Illustration of the surgery costing approach

| **Exemplar patient** | **Surgery**  **(A)** | **Trim-point**  **(B)** | **Length of stay (illustrative)**  **(C)** | **Excess bed days**  **(C-B)** | **Cost of excess bed day cost (£331.99 per day)**  **(D)** | **Stoma management cost (Ileostomy cost per month)**  **(E)** | **Total cost***  **(A+D+E)** |
| --- | --- | --- | --- | --- | --- | --- | --- |
| (i) No complications or other interventions  (HRG FD11F) | £5,363.56 | 10 | 15 | 5 | £1,659.95 | N/A | £7023.51 |
| (ii) No complications or other interventions  (HRG FD11F) - robotic | £5,363.56*1.13  = £6060.82 | 10 | 15 | 5 | £1,659.95 | N/A | £7720.77 |
| (iii) Complication or conversion  (HRG FD11E) | £6,725.82 | 12 | 18 | 6 | £1,991.94 | N/A | £8717.76 |
| (iv) No complication or conversion and stoma  (HRG FD11C) | £7,271.08 | 15 | 18 | 3 | £995.97 | £165.40 | £8,432.45 |
| (v) Complication or conversion and stoma  (HRG FD11B) | £10,432.81 | 23 | 20 | 0 | £0 | £165.40 | £10,598.21 |

HRG FD11F - Malignant Gastrointestinal Tract Disorders with Single Intervention, with CC Score 0-2

HRG FD11E - Malignant Gastrointestinal Tract Disorders with Single Intervention, with CC Score 3-5

HRG FD11C - Malignant Gastrointestinal Tract Disorders with Multiple Interventions, with CC Score 0-2

HRG FD11B - Malignant Gastrointestinal Tract Disorders with Multiple Interventions, with CC Score 3-6

*Plus ICG cost for the ICG+ treatment arm; Plus AL and reintervention costs (details in Table 1A)

Table S2: Unit costs and costing assumptions – procedures and complications

| **Service Type** | **Unit cost** | **Source** | **Notes/Assumptions** |
| --- | --- | --- | --- |
| **Procedure costs** | | | |
| Laparoscopic surgery – no AL, stoma or conversion or complications | £5,363.56 | National schedule of costs FY21-22 | Elective Long stay (EL) Malignant Gastrointestinal Tract Disorders with Single Intervention (with CC Score 0-2)  Bed day trim point = 10 |
| Laparoscopic surgery –complications only (including conversion) | £6,725.82 | National schedule of costs FY21-22 | EL Malignant Gastrointestinal Tract Disorders with Multiple Interventions, with CC Score 3-5  Bed day trim point = 12 |
| Laparoscopic surgery – Stoma no complications | £7,271.08 | National schedule of costs FY21-22 | Malignant Gastrointestinal Tract Disorders with Multiple Interventions (with CC Score 0-2)  Bed day trim point = 15 |
| Laparoscopic surgery – Stoma and complications | £10,432.81 | National schedule of costs FY21-22 | Malignant Gastrointestinal Tract Disorders with Multiple Interventions (with CC Score 3-6)  Bed day trim point = 23 |
| Robotic surgery | N/A | Jayne D, Pigazzi A, Marshall H, Croft J, Corrigan N, Copeland J, Quirke P, West N, Rautio T, Thomassen N, Tilney H, Gudgeon M, Bianchi PP, Edlin R, Hulme C, Brown J. Effect of Robotic-Assisted vs Conventional Laparoscopic Surgery on Risk of Conversion to Open Laparotomy Among Patients Undergoing Resection for Rectal Cancer: The ROLARR Randomized Clinical Trial. JAMA. 2017 Oct 24;318(16):1569-1580 | Costs as above with 1.13 multiplier vs Laparoscopic surgery  Bed days unchanged |
| Excess bed day | £331.98 | For all interventions. 22-23 NT Annex DtA National Tarrif workbook. | Cost of hospital stay per night beyond the trim point for the HRG code. |
| **IFA cost** | | | |
| Contrast cost (Indocyanine green) | £104 | [www.chelwest.nhs.uk/services/support-services/pharmacy/formulary/med-committee/medicines-group-summary-notes-january-2023-4.pdf](https://eur03.safelinks.protection.outlook.com/?url=http%3A%2F%2Fwww.chelwest.nhs.uk%2Fservices%2Fsupport-services%2Fpharmacy%2Fformulary%2Fmed-committee%2Fmedicines-group-summary-notes-january-2023-4.pdf&data=05%7C02%7CD.Meads%40leeds.ac.uk%7C021dedc7c669445569e208dc96f80edd%7Cbdeaeda8c81d45ce863e5232a535b7cb%7C0%7C0%7C638551239840381838%7CUnknown%7CTWFpbGZsb3d8eyJWIjoiMC4wLjAwMDAiLCJQIjoiV2luMzIiLCJBTiI6Ik1haWwiLCJXVCI6Mn0%3D%7C0%7C%7C%7C&sdata=sTqxYB8QTvIuSvetHEJu7JoBy8OvhmPCKsL%2FJzYg%2BYE%3D&reserved=0) |  |
| Cost per equipment use | £44.80 | Liu RQ, Elnahas A, Tang E, Alkhamesi NA, Hawel J, Alnumay A, Schlachta CM. Cost analysis of indocyanine green fluorescence angiography for prevention of anastomotic leakage in colorectal surgery. Surg Endosc. 2022 Dec;36(12):9281-9287 |  |
| **Cost of complications and leak treatment** | | | |
| Pharmacological | £88.31 | BNF  <https://bnf.nice.org.uk/drugs/vancomycin/medicinal-forms/#oral-capsule> | Assume course of Vancomycin 125mg |
| Radiological drain | £4,792.73 | National schedule of costs FY21-22 | Percutaneous single drainage of abdominal abscess with CC score 0-1 YF04C  Bed day trim point = 10 |
| Endoscopy therapy | £2,374.48 | National schedule of costs FY21-22 | FE03A Intermediate therapeutic Endoscopic Upper or lower gastrointestinal tract procedure 19 y and over  Bed day trim point = 0 |
| EUA & Wash out | £1,299.37 | National schedule of costs FY21-22 | Examination under anaesthetic (EUA): For non surgical EUA we assumed a therapeutic colonoscopy and sigmoidoscopy 19 y and over (FE30Z and FE33Z)  Bed day trim point = 0 |
| EUA & Insertion of drain | £1,299.37 | National schedule of costs FY21-22 | Examination under anaesthetic (EUA): For non surgical EUA we assumed a therapeutic colonoscopy and sigmoidoscopy 19 y and over (FE30Z and FE33Z)  Bed day trim point = 0 |
| EUA & re-suturing AL w/o drain | £6,369.60 | National schedule of costs FY21-22 | Weighted average of FD10E, FD10F, FD10G, FD10H. Non-malignant gastrointestinal tract disorder with single intervention cc score 0 to 9+  Bed day trim point = 7 (for FD10H) |
| Laparoscopy & Wash out w/o drain | £6,369.60 | National schedule of costs FY21-22 | Weighted average of FD10E, FD10F, FD10G, FD10H. Non-malignant gastrointestinal tract disorder with single intervention cc score 0 to 9+  Bed day trim point = 7 (for FD10H) |
| Laparoscopy & Wash out w/o drain + dysfunctional stoma | £8,704.60 | National schedule of costs FY21-22 | Weighted average of FD10A, FD10B, FD10C, FD10D. Non-malignant gastrointestinal tract disorder with single intervention cc score 0 to 9+  Bed day trim point = 10 (for FD10D) |
| Laparoscopy & Revision Anastomosis | £6,369.60 | National schedule of costs FY21-22 | Weighted average of FD10E, FD10F, FD10G, FD10H. Non-malignant gastrointestinal tract disorder with single intervention cc score 0 to 9+  Bed day trim point = 7 (for FD10H) |
| Laparotomy & Wash out w/o drain | £6,369.60 | National schedule of costs FY21-22 | Weighted average of FD10E, FD10F, FD10G, FD10H. Non-malignant gastrointestinal tract disorder with single intervention cc score 0 to 9+  Bed day trim point = 7 (for FD10H) |
| Laparotomy & Wash out w/o drain + dysfunctional stoma | £8,704.60 | National schedule of costs FY21-22 | Weighted average of FD10A, FD10B, FD10C, FD10D. Non-malignant gastrointestinal tract disorder with single intervention cc score 0 to 9+  Bed day trim point = 10 (for FD10D) |
| Laparotomy & Revision Anastomosis | £6,369.60 | National schedule of costs FY21-22 | Weighted average of FD10E, FD10F, FD10G, FD10H. Non-malignant gastrointestinal tract disorder with single intervention cc score 0 to 9+  Bed day trim point = 7 (for FD10H) |
| Laparotomy & take down anastomosis | £6,369.60 | National schedule of costs FY21-22 | Weighted average of FD10E, FD10F, FD10G, FD10H. Non-malignant gastrointestinal tract disorder with single intervention cc score 0 to 9+  Bed day trim point = 7 (for FD10H) |
| Stoma removal | £6,369.60 | National schedule of costs FY21-22 | Weighted average of FD10E, FD10F, FD10G, FD10H. Non-malignant gastrointestinal tract disorder with single intervention cc score 0 to 9+  Bed day trim point = 7 (for FD10H) |
| **Other care cost** | | | |
| A+E | £86.88 | NHS reference costs 2021/22  Emergency Medicine, No Investigation with No Significant Treatment, Other type of A&E/minor injury activity with designated accommodation for the reception of accident and emergency patients. | Assumed ratio visit:Tel cost from District nurse |
| A+E (tel/email) | £17.96 | NHS reference costs 2021/22 |  |
| Allied health professional | £174.02 | NHS reference costs 2021/22 | WF01A, Non-Admitted Face-to-Face Attendance, Follow-up, Outpatient ,Consultant Led, Dietetics Service |
| Allied health professional (tel/email) | £35.98 | NHS reference costs 2021/22 | WF01A, Non-Admitted Face-to-Face Attendance, Follow-up, Outpatient ,Consultant Led, Dietetics Service |
| Ambulance | £446.19 | NHS reference costs 2021/22 | See and convey |
| Blood Test | £3.39 | NHS reference costs 2021/22 | DAPS05, Haematology |
| Colonoscopy | £675.25 | NHS reference costs 2021/22 | Therapeutic Colonoscopy, 19 years and over, General Surgery Service |
| CT scan | £103.08 | NHS reference costs 2021/22 | RD20A, Computerised Tomography Scan of One Area, without Contrast, 19 years and over, Imaging: Direct Access |
| Endoscopy | £784.68 | NHS reference costs 2021/22 | Wireless Capsule Endoscopy, 19 years and over  FE50A |
| MRI scan | £178.49 | NHS reference costs 2021/22 | RD01A, Magnetic Resonance Imaging Scan of One Area, without Contrast, 19 years and over, Imaging: Direct Access |
| MRI and sigmoidoscopy | £646.19 | NHS reference costs 2021/22 | Diagnostic Flexible Sigmoidoscopy, 19 years and over, General Surgery Service + MRI |
| PET | £735.35 | NHS reference costs 2021/22 | Positron Emission Tomography with Computed Tomography (PET-CT) of One Area, 19 years and over |
| Radiography | £178.49 | NHS reference costs 2021/22 | Assumed to be MRI scan |
| Specialist nurse | £35.13 | PSSRU 2015 | Assumed same as allied health professional |
| Specialist nurse (tel/email) | £7.26 | Estimated | Assumed same as allied health professional |

Table S3: Unit costs and costing assumptions – patient-reported health care use

| **Service Type** | **Unit cost** | **Source** | **Notes/Assumptions** |
| --- | --- | --- | --- |
| **Secondary care costs** | | | |
| Hospital outpatient clinic (Consultant/Dr) | £188.09 | Unit Costs of Health and Social Care 2022, pg40 | Outpatient, medical specialist palliative care attendance (19 years and over) |
| Hospital outpatient clinic  (Nurse/Other healthcare professional) | £150.70 | Unit Costs of Health and Social Care 2022, pg40 | Outpatient non-medical specialist palliative care attendance |
| Hospital phone contacts (Consultant/Dr) | £21.76 | Unit Costs of Health and Social Care 2022, pg60 | Assumption: BAND 8d; Per surgery consultation lasting 9.22 minutes (PSSRU) |
| Hospital phone contacts (Nurse/Other healthcare professional) | £5.41 | Unit Costs of Health and Social Care 2022, pg73: Nurse-led triage | 5.32 min for telephone consultations |
| Hospital phone contacts (Administrator/Secretary) | £3.56 | Unit Costs of Health and Social Care 2022, pg60 | Assumption based on 5 minutes for Band 3 (2+ yrs) |
| **Primary, community and social care costs** | | | |
| Doctor GP surgery (visit) | £46.45 | Unit Costs of Health and Social Care 2022, pg70 | Per surgery consultation lasting 9.22 minutes  - including direct care, with qualification costs |
| Doctor GP surgery(tel/email) | £27.21 | Unit Costs of Health and Social Care 2022, pg70 & Clinical workload in UK primary care: a retrospective analysis of 100 million consultations in England, 2007–14 Hobbs R 2016 The lancet 387, 10035 | Assumption: consultation lasting 5.4 minutes, based on £41 per 9.22 minutes consultation (cost per minute £4.45. Same cost for e-mail consultation) |
| Doctor GP surgery (home visit) | £117.90 | Unit Costs of Health and Social Care 2015, pg176 & Unit Costs of Health and Social Care 2022, pg70 | Home visit duration 11.4 minutes plus 12 minutes travel time. We assume travel time at not f2f contact rate - including direct care, with qualification costs |
| GP administrator/receptionist | £3.56 | Unit Costs of Health and Social Care 2022, pg60 | Assumption based on 5 minutes for Band 3 (2+ yrs) |
| Practice nurse | £19.79 | Unit Costs of Health and Social Care 2022, pg68 | £52 per hour with qualifications. Assumption: Per surgery consultation lasting 15.5 minutes. (from PSSRU 2015) - including direct care, with qualification costs. |
| Practice nurse (tel/email) | £7.26 | Unit Costs of Health and Social Care 2022, pg68 & Clinical workload in UK primary care: a retrospective analysis of 100 million consultations in England, 2007–14 Hobbs R 2016 The lancet 387, 10035 | Assumption: consultation lasting 5.69 minutes assuming a ratio of 1:030 f2f contact. We assumed that e-mail consultation has the same cost as telephone consultation |
| District nurse (home) | £40.58 | PSSRU 2015 | Assumes district nurse will only visit patients homes: based on PSRRU 2015 15.5 minutes per vist at £67 per hour of patient related work, plus travel time assumed at 12 minutes |
| District nurse (tel/email) | £7.20 | Unit Costs of Health and Social Care 2022, pg68 & Clinical workload in UK primary care: a retrospective analysis of 100 million consultations in England, 2007–14 Hobbs R 2016 The lancet 387, 10035 | 5.69 minutes per consultation assuming same cost as f2f patient related work |
| NHS 24 or NHS 111 | £13.14 | Turner J, Knowles E, Simpson R, Sampson F, Dixon S, Long J, et al. Impact of NHS 111 Online on the NHS 111 telephone service and urgent care system: a mixed-methods study. Health Serv Deliv Res 2021;9(21). | Inflated from £11.40 in 2021. |
| Out of hours GP | £94.50 | Department of Health and NHS England, Out of hours GP services in England | Based on a cost of £68.30 (2013-14) from Out of Hours GP service in England pg 15.  - including direct care, with qualification costs |
| Out of hours GP (tel/email) | £17.56 | Unit Costs of Health and Social Care 2022, pg73 GP-led triage | Cost of telephone triage by a GP |
| Psychiatrist/Psychologist/  Counsellor | £67.99 | Unit Costs of Health and Social Care 2022, pg60 | Assumption Band 6: 1 hour ratio 1:091 (page 58) |
| Psychiatrist/Psychologist/  Counsellor (home) | £81.59 | Unit Costs of Health and Social Care 2022, pg60 | Assumption: One hour consultation plus 12 minutes travel time at f2f cost |
| Psychiatrist/Psychologist/  Counsellor (tel/email) | £10.45 | Unit Costs of Health and Social Care 2022, pg60 | Assumption Band 6: 1 hour as a f2f consultation Ratio 1:091 Assuming same duration as specialist consultation 9.22 minutes |
| Physiotherapist | £163.16 | Unit Costs of Health and Social Care 2022, pg40 | Average cost per session (one-to-one) |
| Physiotherapist (home) | £195.79 | Unit Costs of Health and Social Care 2022, pg40 | Assumption: average cost per session (one-to-one) plus travel time 12 minutes |
| Physiotherapist (tel/email) | £15.47 | Unit Costs of Health and Social Care 2022, pg40 | Assumption: 5.69 min for telephone consultations (£144/60*5.69) |
| Dietitian | £113.30 | Unit Costs of Health and Social Care 2022, pg40 | Average cost per session (one-to-one) |
| Dietitian (home) | £135.97 | Unit Costs of Health and Social Care 2022, pg40 | Average cost per session (one-to-one), plus 12 minutes transfer time |
| Dietitian (tel/email) | £37.77 | Unit Costs of Health and Social Care 2022, pg40 | Assumption: 20 min for telephone consultations (£100/60*20) |
| Occupational therapist | £133.70 | Unit Costs of Health and Social Care 2022, pg40 | Average cost per session (one-to-one) |
| Occupational therapist (home) | £160.44 | Unit Costs of Health and Social Care 2022, pg40 | Average cost per session (one-to-one) plus 12 minutes transfer time |
| Occupational therapist (tel/email) | £11.94 | Unit Costs of Health and Social Care 2022, pg40 | Assumption: 5.32 min for telephone consultations (£118/60*5.32) |

Table S4: Impact of ALs on Costs and HRQoL

| **Parameter** | **N** | **Mean impact of AL** | **P value** | **Lower CI** | **Upper CI** |
| --- | --- | --- | --- | --- | --- |
| **Procedural resource use** |  |  |  |  |  |
| Time in theatre (minutes) | 324 | 41.84 | **0.013** | 8.86 | 74.81 |
| Length of stay (days) | 342 | 7.85 | **0.000** | 4.72 | 10.31 |
| **Costs*** |  |  |  |  |  |
| Total costs – 90 day | 93 | £4330.07 | **0.001** | £1683.88 | £6976.27 |
| Surgery costs – 90 day | 285 | £878.58 | 0.351 | -£967.31 | £2724.48 |
| Inpatient costs – 90 day | 142 | £1086.04 | 0.338 | -£1135.14 | £3307.22 |
| Outpatient costs – 90 day | 155 | -£290.80 | 0.411 | -£984.30 | £402.70 |
| Primary care costs – 90 day | 153 | -£16.79 | 0.789 | -£139.64 | £106.07 |
| Medication costs – 90 day | 110 | £27.12 | 0.609 | -£76.68 | £130.92 |
| **HRQoL** |  |  |  |  |  |
| EQ-5D-5L - 90 day | 255 | -0.084 | **0.011** | -0.149 | -0.019 |
| EQ-5D-5L – 12 month | 199 | -0.046 | 0.297 | -0.134 | 0.041 |
| **QALYs** |  |  |  |  |  |
| QALYs – 90 day | 255 | -0.013 | **0.023** | -0.024 | -0.001 |
| QALYs – 12 month | 194 | -0.066 | 0.052 | -0.133 | 0.001 |
| **Net Health Benefit**** |  |  |  |  |  |
| NHB – 90 day | 93 | -0.257 | **0.000** | -0.401 | -0.113 |

*Not all categories of cost are included (e.g. cost of diagnosing and treating ALs); **Willingness to pay per QALY = £20,000

Figure S1 Cost-effectiveness acceptability curve

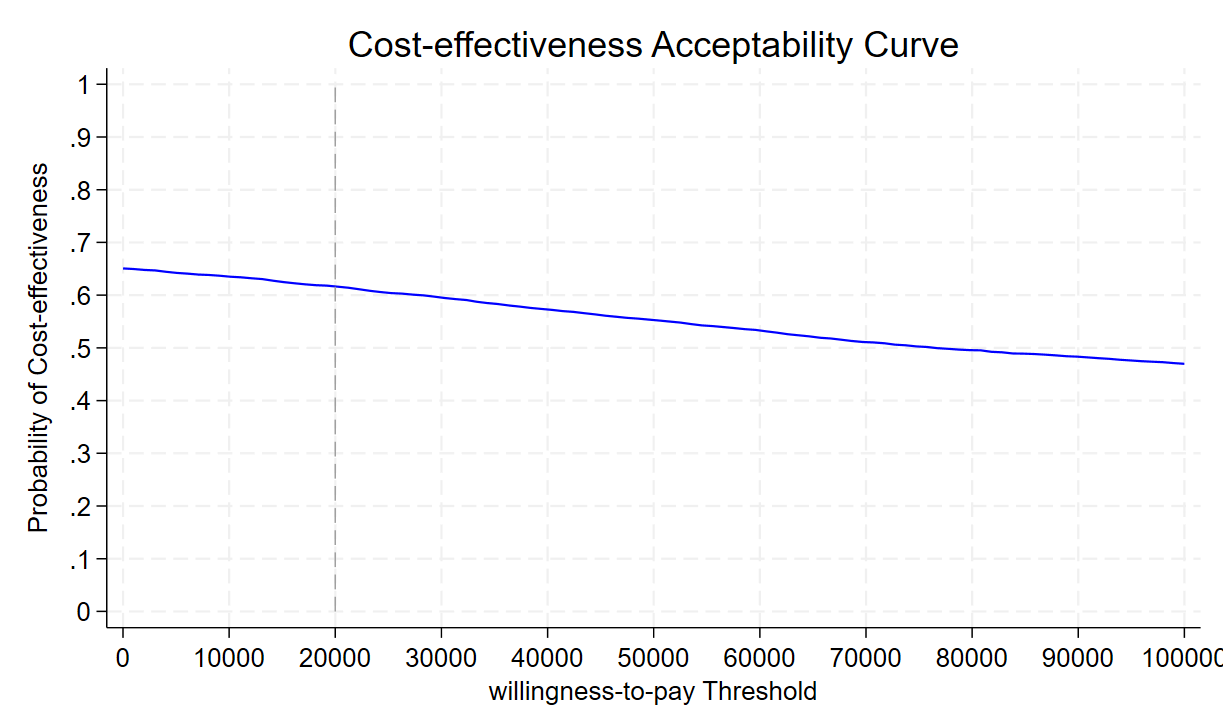
